# Improvements in Patient-Reported Functioning after Lung Transplant is Associated with Improved Quality of Life and Survival

**DOI:** 10.1101/2024.08.05.24311510

**Authors:** Leslie Seijo, Ying Gao, Legna Betancourt, Aida Venado, Steven R. Hays, Jasleen Kukreja, Danny R. Calabrese, John R. Greenland, Jonathan P. Singer

**Author notes:** Corresponding Author X Handle: @lseijoMD, Business Address and Reprints Requests: University of California, San Francisco 505 Parnassus Ave, Suite M1083B Business. **Funding:** JPS: NHLBI K23HL111115, U01HL163242, U01HL145435. **Author Contributions:** All authors listed above agree to be accountable for all aspects of the work in ensuring that questions related to the accuracy or integrity of any part of the work are appropriately investigated and resolved. The authors made the following contributions - LS, JPS, and YG made substantial contributions to the conception and study design. - JPS acquired the research funding for the study. - LS, JPS, YG, and LB made substantial contributions to the acquisition, analysis, or interpretation of data for the work. - LS and JPS wrote the first draft of the manuscript. All authors revised the manuscript for important intellectual content. - All authors approved the manuscript. Author Conflicts of Interest: - JPS: Consulting fees from XVIVO; Scientific Advisory Board: Mallinckrodt Pharmaceuticals; DSMB: Krystal Biotech - SRH: Consulting fees from AI Therapeutics and CareDx; Scientific Advisory Board: CareDx - JK: DSMB Lung Bioengineering - JRG: Scientific Advisory Board and Research Funding: Theravance Biopharma Mallinckrodt Pharmaceuticals.

## Abstract

Lung transplantation aims to improve health-related quality of life (HRQL) and survival. While lung function improvements are associated with these outcomes, the association between physical functioning and these outcomes is less clear. We investigated the association between changes in patient-reported physical functioning and HRQL, chronic lung allograft dysfunction (CLAD), and survival after lung transplantation.

This single-center prospective cohort study analyzed 220 lung transplant recipients who completed the 15-item Lung Transplant Valued Life Activities (LT-VLA) before and repeatedly after transplant. HRQL was assessed using generic, respiratory disease-specific, and utility measures. Associations between 0.3-point changes (the minimally important difference) in LT-VLA as a time-varying predictor on HRQL, CLAD, and mortality were tested using linear regression and Cox proportional hazard models. Models were adjusted for demographics, disease diagnosis, and post-operative lung function as a time-varying covariate.

Participants were 45% female and 75% White, with a mean age of 56 (±12) years. Each 0.3-point improvement in LT-VLA was associated with substantially improved HRQL across all measures (adjusted p-values <0.01). Each 0.3-point improvement in LT-VLA was associated with a 13% reduced hazard of CLAD (adjusted HR: 0.87, 95% CI: 0.76–0.99, p=0.03) and a 19% reduced hazard of mortality (adjusted HR: 0.81, 95% CI: 0.67–0.95, p=0.01).

Improvements in patient-reported physical functioning after lung transplantation are associated with improved HRQL and reduced risk of CLAD and death, independent of allograft function. The simplicity of the LT-VLA suggests it could be a valuable monitoring or outcome measure in both clinical and research settings.

## Introduction

Since its inception, the clinical aims of lung transplantation have been to improve respiratory failure-attributable physical impairments, improve health-related quality of life (HRQL), and extend survival.^1,2^ Yet, despite advancements in surgical techniques and post-operative care, a disparity in outcomes persists, leaving some recipients with enduring disability and compromised HRQL.^3,4^ Identifying modifiable factors that influence outcomes in lung transplantation remains a significant challenge.^5,6^

Most lung-transplant-related research has appropriately focused on graft-related complications, given their association with death after transplant.^7^ Emerging lines of research are focused on physical functioning as a key determinant of patient-centered outcomes (PCO) after lung transplant.^4,8^ In conditions such as chronic obstructive pulmonary disease, impairments in physical function are associated with worse HRQL and mortality.^9,10^ However, while informative, conventional measures used to assess physical functioning such as actigraphy, lab-based measures of exercise capacity (e.g., six-minute walk distance),^11^ or surveys assessing activities of daily living (ADL)^4,12^ are either cumbersome to utilize frequently or, in the case of ADLs, fail to detect the early onset of impairments in physical functioning that can precede more evident daily limitations. To address this gap, the Lung Transplant-Valued Life Activities (LT-VLA) scale was developed. The LT-VLA is a unidimensional patient-reported physical functioning/disability measure developed and validated for use in advanced lung disease and lung transplantation.^13^

In this study, we investigated whether improvements in patient-reported physical functioning and disability are associated with improved outcomes after lung transplantation. We specifically investigated whether improved physical functioning was associated with enhanced HRQL as well as reduced risk of chronic lung allograft dysfunction (CLAD) and mortality after lung transplantation.

## Materials and methods

### Study design, participants, and setting

We analyzed data from the “Breathe Again” cohort at the University of California San Francisco (UCSF), a prospective study examining the impact of lung transplantation on a range of patient-centered outcomes. Breathe Again enrolled adults 18 years of age or older undergoing their first lung transplant between February 2010 to January 2017.^12^

Participants were enrolled near the time of placement on the lung transplant waiting list. At enrollment, a multi-instrument patient-reported outcomes (PRO) battery was administered. While on the waitlist for a lung transplant, assessments were repeated every three months or when there were major significant clinical changes such as hospitalization. After the transplantation, the same PRO battery was performed at 3, 6, 12, 18, and 24 months. Clinical and demographic variables were collected from the electronic medical record. All study participants provided written informed consent. The UCSF Committee on Human Research approved the study. The study complied with the ISHLT Ethics statement.

### Conceptual Model

Our study is framed by a conceptual model of disablement developed by Nagi^15^ and later adopted by the Institute of Medicine to examine the continuum from disease pathology to its impact on daily life and, subsequently, HRQL and mortality.^15^ In this model, disease results in impaired organ functioning. This impairment leads to functional limitations, which can be quantified in a laboratory-based setting (e.g., pulmonary function testing or six-minute walk distance). Functional limitations, in turn, result in disability, defined as limitations in performing activities in daily life. Within this theoretical framework, functional limitations and disability are considered precursors to both HRQL and mortality.^16^ For this analysis, we hypothesized that improvements in patient-reported physical functioning/disability would be associated with improved HRQL and survival (Figure 1). Since physical functioning can reflect health status beyond lung function, we further hypothesized that improvements in physical disability might be associated with HRQL and survival even after accounting for allograft function. Given our prior observation that frailty onset was associated with worse LT-VLA disability as well as increased risk of subsequent CLAD, we also explored whether improvement in LT-VLA was a marker for reduced risk of CLAD.^17,18^

**Figure 1:**
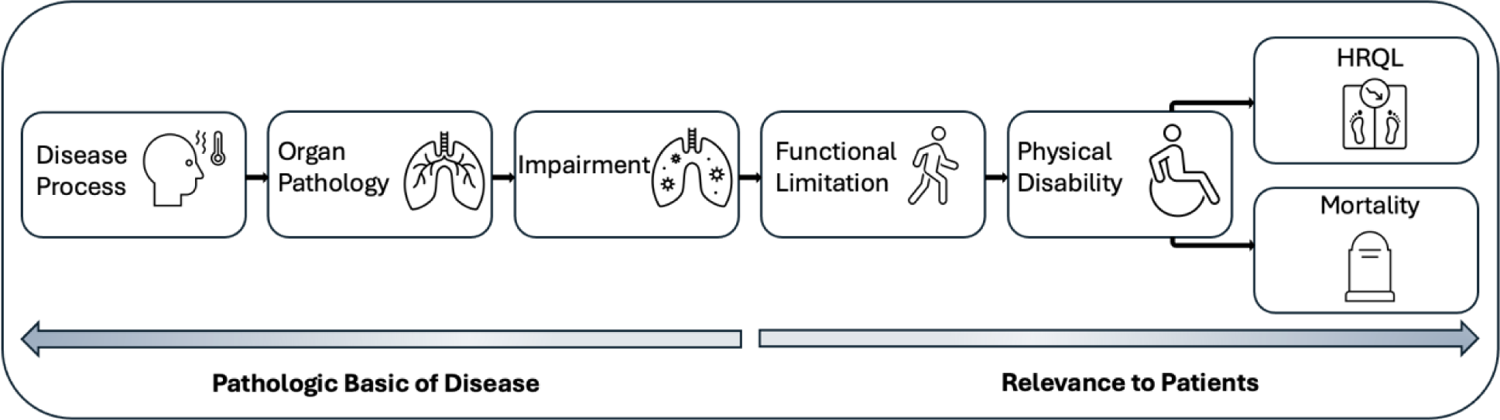
Conceptual model of disablement. Proposed by Nagi^15^, illustrates the progression from disease process to disability and its consequent effect on health-related quality of life and mortality.

### Exposure Variables

Lung Transplant Valued Life Activities (LT-VLA) scale: Patient-reported physical functioning and disability were assessed using the LT-VLA. The LT-VLA is a 15-item self-rated instrument of physical disability designed and validated for patients with advanced lung disease and lung transplantation.^13^ The LT-VLA queries how much difficulty respondents have in completing physical activities spanning three hierarchical domains: those activities essential for self-care; those activities central to one’s principal social role(s) such as employment or schooling; and those activities that enhance enjoyment and life satisfaction, such as the ability to travel or visit friends and family.

For each of the 15 items, participants rate how much difficulty they have in performing each activity on a 4-point scale, ranging from 0 (no difficulty) to 3 (unable to perform), with an option for ‘does not apply,’ ensuring relevance to the participant’s lifestyle.^13^ The aggregate mean score, calculated as the sum of rated items divided by the number of items applicable to the participant, spans from 0 to 3, in which lower scores indicate improvements in physical functioning. Prior work has demonstrated a change of 0.3 meets the minimally important difference.^13^

### Outcome Variables

HRQL: HRQL is inherently multi-dimensional, encompassing various dimensions pertinent to lung transplant recipients.^8^ To comprehensively evaluate HRQL, we employed a combination of generic and disease-specific assessment tools. The Medical Outcomes Study Short Form Physical and Mental Component Summary Scales (SF-12 PCS and MCS) provided a broad measure of generic HRQL.^19,20^ These scales span from 0 to 100 points, with higher scores indicating better HRQL. A change of five points is considered clinically significant.^19,20^ Respiratory specific-HRQL was quantified using the Airway Questionnaire 20-Revised (AQ20-R) (range 0-20, lower scores represent better HRQL, and a change of 1.75 points meets the minimally important difference).^21,22^ Health utilities were assessed with the EuroQol 5D (EQ5D; range −0.11-1; higher scores denote improved health utilities; a change of 0.06 meets the minimally important difference).^23^

#### CLAD

CLAD was defined as a 20% decline in forced expiratory volume in one second (FEV1) from the post-lung transplant baseline that persisted for at least 3 months.^24^ Time to CLAD was calculated as the number of days from the date of transplant to the date of definite CLAD onset.

#### Survival

Survival time was determined as the number of days from the date of transplant to the date of death. For this analysis, administrative censorship was applied at 48 months post-lung transplant.

### Confounding and precision variables

Clinical demographics and measures such as age, sex, diagnosis, body mass index (BMI, kg/m2), FEV1 (measured in liters), and forced vital capacity (FVC, measured in liters) were abstracted from electronic medical records. Post-lung transplant, we obtained FEV1, FVC, and BMI data from the clinical visits coinciding with our research study visits. Of the repeated pre-lung transplant assessment, we used the one collected most proximal to the date of transplant as the pre-transplant baseline assessment.

### Analytic approach

We tested the association between LT-VLA and HRQL by linear mixed effects models for each HRQL measure. LT-VLA was modeled as a time-varying predictor, allowing us to evaluate how changes in LT-VLA scores influenced HRQL over time. ^25^ All models were adjusted for age, sex, race, and diagnostic indication for lung transplant. As a secondary analysis, post-lung transplant FEV1 was included as a time-dependent covariate allowing us to estimate the effect of changes in LT-VLA on HRQL independent of allograft function. For CLAD, Cox proportional hazard models were employed, modeling LT-VLA as a time-varying predictor. This approach enabled us to examine the impact of changes in LT-VLA on the *subsequent* risk of developing CLAD. For mortality, we also utilized Cox proportional hazard models with LT-VLA as a time-varying predictor. Our primary models were adjusted for demographic factors. A secondary analysis included post-lung transplant FEV1 as a time-dependent covariate to test the effect of changes in LT-VLA on mortality risk independent of allograft function.

Since not all participants had completed 3-years of study visits when Breathe Again ended, the decision of when to administratively censor follow-up time for CLAD and mortality based on external literature was not clear. Thus, we elected to consider two different time points for administrative censoring for these outcomes. For our primary analyses, we censored follow-up time at one year after the last LT-VLA measure for mortality and two years after the last LT-VLA measure for CLAD. As sensitivity analyses, we censored follow-up time at four years post-transplant for death and five years post-transplant for CLAD. These sensitivity analyses were performed to challenge our initial modeling decisions and verify the consistency of our results over longer follow-up periods. Statistical analyses were performed using SAS (version 9.4, SAS Institute, Cary, North Carolina) and R (version 4.0.5, R Foundation for Statistical Computing, Vienna, Austria).

## Results

The study included 220 lung transplant recipients with a mean age of 56 ± 12 years (Table 1). The cohort consisted of 99 females (45%). In terms of race and ethnicity, 164 (75%) were White, 16 (7%) were Black, 11 (5%) were Asian, and 29 (13%) were Hispanic. Most participants underwent lung transplantation for pulmonary fibrosis (72%) followed by non-suppurative obstructive lung diseases (16%). Baseline HRQL measures included the SF12PCS with a median score of 23.6 (IQR: 18.6, 28.3), AQ20R with a median score of 13.0 (IQR: 11.0, 16.0), and EQ5D with a median score of 0.7 (IQR: 0.6, 0.8).

**Table 1:**
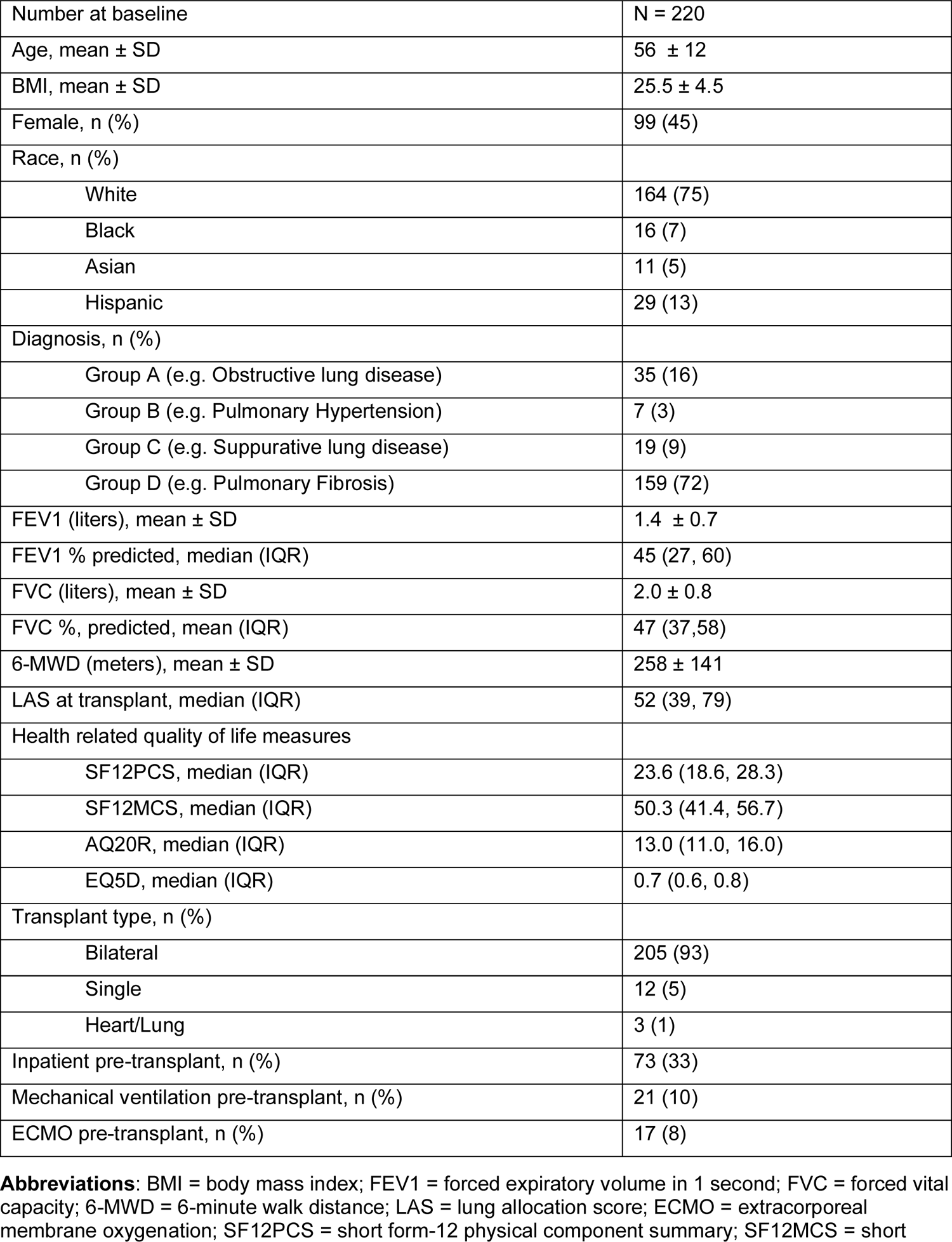
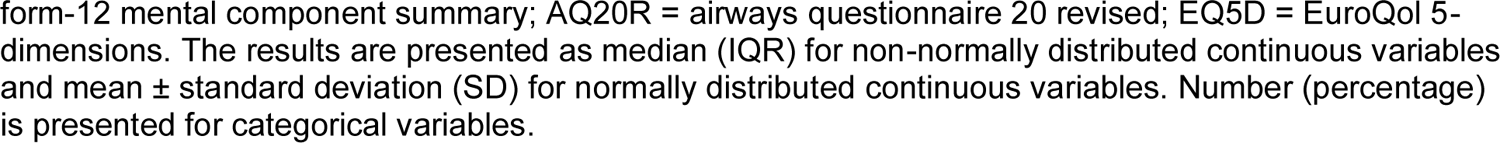
Baseline Demographics and Clinical Characteristics of Lung Transplant Recipients (N = 220)

|  |  |
| --- | --- |
| Number at baseline | N = 220 |
| Age, mean ± SD | 56 ± 12 |
| BMI, mean ± SD | 25.5 ± 4.5 |
| Female, n (%) | 99 (45) |
| Race, n (%) |  |
| White | 164 (75) |
| Black | 16 (7) |
| Asian | 11 (5) |
| Hispanic | 29 (13) |
| Diagnosis, n (%) |  |
| Group A (e.g. Obstructive lung disease) | 35 (16) |
| Group B (e.g. Pulmonary Hypertension) | 7 (3) |
| Group C (e.g. Suppurative lung disease) | 19 (9) |
| Group D (e.g. Pulmonary Fibrosis) | 159 (72) |
| FEV1 (liters), mean ± SD | 1.4 ± 0.7 |
| FEV1 % predicted, median (IQR) | 45 (27, 60) |
| FVC (liters), mean ± SD | 2.0 ± 0.8 |
| FVC %, predicted, mean (IQR) | 47 (37,58) |
| 6-MWD (meters), mean ± SD | 258 ± 141 |
| LAS at transplant, median (IQR) | 52 (39, 79) |
| Health related quality of life measures |  |
| SF12PCS, median (IQR) | 23.6 (18.6, 28.3) |
| SF12MCS, median (IQR) | 50.3 (41.4, 56.7) |
| AQ20R, median (IQR) | 13.0 (11.0, 16.0) |
| EQ5D, median (IQR) | 0.7 (0.6, 0.8) |
| Transplant type, n (%) |  |
| Bilateral | 205 (93) |
| Single | 12 (5) |
| Heart/Lung | 3 (1) |
| Inpatient pre-transplant, n (%) | 73 (33) |
| Mechanical ventilation pre-transplant, n (%) | 21 (10) |
| ECMO pre-transplant, n (%) | 17 (8) |

**Abbreviations:** BMI = body mass index; FEV1 = forced expiratory volume in 1 second; FVC = forced vital capacity; 6-MWD = 6-minute walk distance; LAS = lung allocation score; ECMO = extracorporeal membrane oxygenation; SF12PCS = short form-12 physical component summary; SF12MCS = short
form-12 mental component summary; AQ20R = airways questionnaire 20 revised; EQ5D = EuroQol 5-dimensions. The results are presented as median (IQR) for non-normally distributed continuous variables and mean ± standard deviation (SD) for normally distributed continuous variables. Number (percentage) is presented for categorical variables.

### Association of LT-VLA with Health-Related Quality of Life

Improvement in LT-VLA was strongly associated with improved HRQL. A 0.3 unit increase in LT-VLA was associated with a 4-point improvement in SF12PCS scores in both unadjusted (Estimate: 4.15, 95% CI: 3.79–4.51, p < 0.0001) and adjusted models (Estimate: 4.15, 95% CI: 3.79–4.51, p < 0.0001). Further adjusting for post-operative allograft function only modestly attenuated the strength of the association (Estimate: 3.90, 95% CI: 3.54–4.26, p < 0.0001). Similar positive associations were observed for the SF12MCS, AQ20R, and EQ5D measures, indicating that improvements in physical functioning are associated with better HRQL and health utilities (Table 2).

**Table 2:** Association of LT-VLA with Health-Related Quality of Life.

| Outcome Variable | Conceptual Domain | Model | Parameter Estimate (95% CI) | P Value |
| --- | --- | --- | --- | --- |
| **SF12PCS**<br>(MCID = 5) | Generic physical HRQL | Unadjusted | 4.15 (3.79, 4.51) | <0.0001 |
|  |  | Adjusted for demographics | 4.15 (3.79, 4.51) | <0.0001 |
|  |  | Adjusted for demographics & lung function | 3.90 (3.54, 4.26) | <0.0001 |
| **SF12MCS**<br>(MCID = 5) | Generic mental HRQL | Unadjusted | 1.98 (1.64, 2.32) | <0.0001 |
|  |  | Adjusted for demographics | 1.98 (1.64, 2.33) | <0.0001 |
|  |  | Adjusted for demographics & lung function | 1.99 (1.62, 2.35) | <0.0001 |
| **AQ20R**<br>(MCID = 1.75) | Respiratory-specific HRQL | Unadjusted | -0.85 (-0.98, -0.73) | <0.0001 |
|  |  | Adjusted for demographics | -0.87 (-0.99, -0.74) | <0.0001 |
|  |  | Adjusted for demographics & lung function | -0.76 (-0.89, -0.64) | <0.0001 |
| **EQ5D**<br>(MCID = 0.06) | Health Utility | Unadjusted | 0.06 (0.05, 0.06) | <0.0001 |
|  |  | Adjusted for demographics | 0.06 (0.05, 0.06) | <0.0001 |
|  |  | Adjusted for demographics & lung function | 0.06 (0.05, 0.06) | <0.0001 |

**Abbreviations:** LT-VLA = Lung Transplant-Valued Life Activities; CI = Confidence Interval; SF12PCS = Medical Outcomes Study Short Form 12 Physical Component Summary Scale; SF12MCS = Medical Outcomes Study Short Form 12 Mental Component Summary Scale; AQ20R = Airway Questionnaire 20-Revised; EQ5D = Euroqol 5D.
**Note:** Parameter estimate represents changes in PRO per 0.3 improvement in LT-VLA using linear mixed effect modeling with LT-VLA as a time-varying predictor. All models were adjusted for age, sex, race, and diagnostic indication for lung transplant. The adjusted models for demographics and lung function include post-lung transplant FEV1 as a time-dependent covariate.

### Association of LT-VLA with Chronic Lung Allograft Dysfunction

Improvements in LT-VLA were associated with a decreased risk of CLAD. In the unadjusted model, a 0.3-unit improvement in LT-VLA was associated with a trend toward reduced risk of CLAD (HR: 0.89, 95% CI: 0.78–1.01, p = 0.063). After adjusting for demographics, improvement in LT-VLA was associated with a 13% lower hazard of subsequent CLAD (HR: 0.87, 95% CI: 0.76–0.99, p = 0.033). A sensitivity analysis extending time under observation to five years post-lung transplant was consistent with our primary findings. Improved LT-VLA was associated with reduced CLAD risk in both unadjusted (HR: 0.89, 95% CI: 0.79–1.00, p = 0.05) and adjusted models (HR: 0.87, 95% CI: 0.77–0.98, p = 0.033) (Table 3).

**Table 3:**
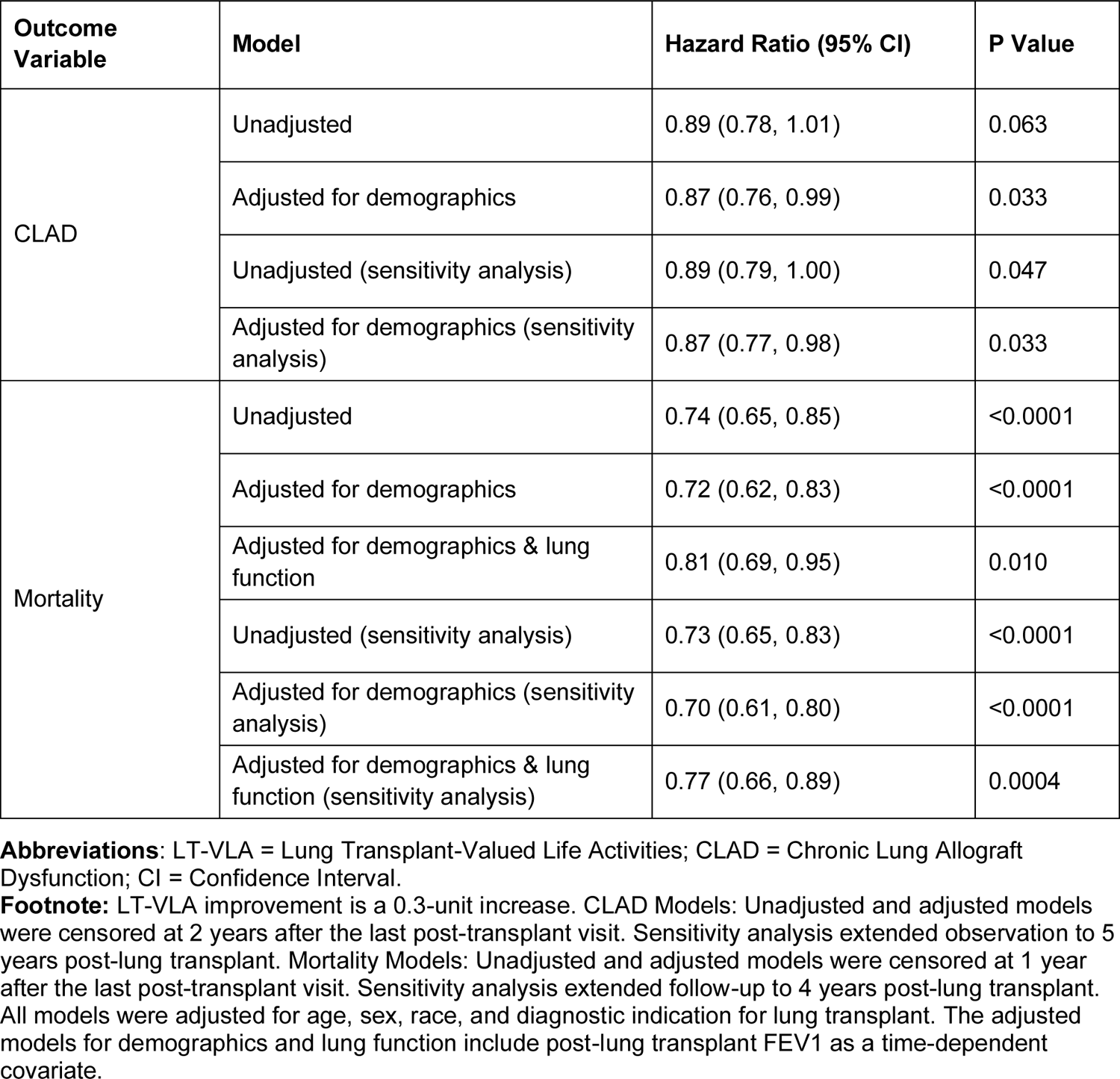
Association of LT-VLA with CLAD and Mortality.

| Outcome Variable | Model | Hazard Ratio (95% CI) | P Value |
| --- | --- | --- | --- |
| CLAD | Unadjusted | 0.89 (0.78, 1.01) | 0.063 |
|  | Adjusted for demographics | 0.87 (0.76, 0.99) | 0.033 |
|  | Unadjusted (sensitivity analysis) | 0.89 (0.79, 1.00) | 0.047 |
|  | Adjusted for demographics (sensitivity analysis) | 0.87 (0.77, 0.98) | 0.033 |
| Mortality | Unadjusted | 0.74 (0.65, 0.85) | <0.0001 |
|  | Adjusted for demographics | 0.72 (0.62, 0.83) | <0.0001 |
|  | Adjusted for demographics & lung function | 0.81 (0.69, 0.95) | 0.010 |
|  | Unadjusted (sensitivity analysis) | 0.73 (0.65, 0.83) | <0.0001 |
|  | Adjusted for demographics (sensitivity analysis) | 0.70 (0.61, 0.80) | <0.0001 |
|  | Adjusted for demographics & lung function (sensitivity analysis) | 0.77 (0.66, 0.89) | 0.0004 |

**Abbreviations:** LT-VLA = Lung Transplant-Valued Life Activities; CLAD = Chronic Lung Allograft Dysfunction; CI = Confidence Interval.
**Footnote:** LT-VLA improvement is a 0.3-unit increase. CLAD Models: Unadjusted and adjusted models were censored at 2 years after the last post-transplant visit. Sensitivity analysis extended observation to 5 years post-lung transplant. Mortality Models: Unadjusted and adjusted models were censored at 1 year after the last post-transplant visit. Sensitivity analysis extended follow-up to 4 years post-lung transplant. All models were adjusted for age, sex, race, and diagnostic indication for lung transplant. The adjusted models for demographics and lung function include post-lung transplant FEV1 as a time-dependent covariate.

### Association of LT-VLA with Mortality

Improvements in LT-VLA were associated with reduced mortality. A 0.3-unit improvement in LT-VLA was associated with a 26% reduction in mortality risk in unadjusted models (HR: 0.74, 95% CI: 0.65–0.85, p < 0.0001). This association was similar after adjusting for demographics (aHR: 0.72, 95% CI: 0.62–0.83, p < 0.0001) and for both demographic factors and post-operative allograft function (aHR: 0.81, 95% CI: 0.69–0.95, p = 0.010) (Table 3). A sensitivity analysis extending follow-up time to four years post-transplant was consistent with our primary findings. Improvements in LT-VLA were associated with improved survival in unadjusted (HR: 0.73, 95% CI: 0.65–0.83, p < 0.0001) and adjusted models (adjusted for demographics HR: 0.70, 95% CI: 0.61–0.80, p < 0.0001; adjusted for demographics and allograft function. HR: 0.77, 95% CI: 0.66–0.89, p < 0.0001) (Table 3).

## Discussion

This study demonstrates that improvements in patient-reported physical functioning up to 3 years after lung transplant are strongly associated with enhanced HRQL as well as reduced risk of CLAD and mortality. Further, that improvements in LT-VLA were associated with reduced risk of mortality independent of allograft function highlights the importance of patient-reported physical functioning/disability as an important factor in outcomes after lung transplantation. Our findings suggest that interventions to improve at improving physical functioning may have significant clinical benefits across the range of outcomes important to the field of lung transplantation.

The 15-item LT-VLA scale requires 2-3 minutes to complete and offers a unique and patient-centered approach to assessing physical functioning and disability in lung transplant recipients.^13^ Unlike traditional measures such as the six-minute walk distance (6-MWD), actigraphy-based assessments of physical activity, or (instrumental) activities of daily living, the LT-VLA captures a person’s perceived difficulty in performing a range of physical activities that are personally significant to them.^13^ This distinction makes the LT-VLA a valuable, portable, and easily scalable complement to these traditional tools, providing a comprehensive view of the functional status and disability of lung transplant recipients.

That changes in LT-VLA scores were associated with enhanced HRQL, reduced CLAD risk, and decreased mortality, underscores the clinical and research value of measures directly assessed from patients. LT-VLA scores were correlated with improved HRQL, highlighting the broader impact of physical functioning on overall well-being in lung transplant recipients. If confirmed, the association between LT-VLA and CLAD and mortality risk suggests that LT-VLA may serve as a valuable prognostic tool for identifying patients at higher risk for poor outcomes. It also may identify those who could benefit from targeted interventions.

Our findings align with previous studies that have highlighted the importance of physical functioning in predicting outcomes in other chronic lung diseases. Studies in chronic obstructive pulmonary disease have shown that reduced physical functioning is associated with worse HRQL and increased mortality.^26,27^ Similarly, work in idiopathic pulmonary fibrosis has demonstrated the prognostic value of physical functioning measures, such as the 6-MWD and gait speed, in predicting patient outcomes like hospitalization and mortality.^28–30^ In the context of lung transplantation, previous studies have examined the prognostic value of the Karnofsky Performance Status (KPS) scale, a clinician-rated assessment of overall global health developed for oncology patients, showing its association with mortality post-lung transplantation.^2,31^ However, KPS evaluates overall health rather than specific physical functioning and is not patient-reported. Our study builds on these lines of research in the broader chronic lung disease literature by demonstrating the prognostic value of patient-reported improvements in physical functioning.

Although our findings are consistent with Nagi’s conceptual model of disablement, the mechanisms underpinning our observations are unknown. Based on our conceptual model, a possible mechanism that might explain the associations between LT-VLA with HRQL, CLAD, and mortality may be that physical functioning reflects overall health status, including factors such as muscle strength, endurance, and frailty. Impairments in these areas can lead to negative health events, ultimately affecting survival and quality of life.^11,32^ Frailty onset after lung transplant has been linked to worse HRQL, onset of CLAD, and mortality.^17,33^ Therefore, LT-VLA could serve as a valuable marker for improving frailty and overall patient outcomes.

Our findings have implications for both clinical practice and research. Worsening of LT-VLA scores in clinical practice could help identify patients at higher risk for poor outcomes and potentially tailor care plans to address the root causes of LT-VLA declines. For example, patients showing declines in LT-VLA scores might benefit from pulmonary rehabilitation,^34,35^ addressing causes for declines in exercise (e.g., depression, pain, weight gain), or closer clinical monitoring.^36^ Additionally, the LT-VLA could be integrated into remote-based research programs and remote monitoring initiatives, facilitating continuous assessment and management of patient health outside of clinical settings. Further, the LT-VLA may be a simple, informative, patient-reported outcome for interventions targeting physical functioning. This aligns with the growing trend towards telemedicine and remote patient monitoring in research, which has been shown to improve patient outcomes and reduce healthcare costs.^11,37^

Our study has several strengths. First, it utilizes a patient-centered measure, the LT-VLA scale, which captures disability and physical functioning from the perspective of lung transplant recipients. Our longitudinal study design allowed us to identify novel associations between changes in physical functioning on several key clinical outcomes. Our use of validated measures and outcomes and adjustment for important covariates enhance the rigor of our findings.

Despite its strengths, our study has limitations. The single-center design may limit the generalizability of our findings to other populations and settings. The observational nature of the study precludes establishing causal relationships between LT-VLA scores and clinical outcomes. The reliance on self-reporting for the LT-VLA may introduce reporting biases and variability seen in other PROs. Although we adjusted for key demographic and clinical variables, there may be residual confounding factors that were not accounted for in our analyses. Potential residual confounders could include the participants’ socioeconomic status, which can influence access to healthcare and post-lung transplant care, and the level of social support, which can impact physical functioning and overall recovery. Psychological factors such as depression and anxiety, which are known to affect HRQL and physical functioning, were not fully accounted for and may have influenced the outcomes.^38,39^ Future studies should aim to measure and adjust for these factors to provide a more comprehensive understanding of the associations between LT-VLA scores and clinical outcomes.

Ongoing research is focused on validating the LT-VLA scale in a large, multi-center cohort to confirm the generalizability of our findings. Randomized controlled trials are needed to establish causal relationships between improvements in LT-VLA scores and clinical outcomes. Research should also explore the integration of LT-VLA monitoring into routine clinical practice and remote monitoring programs, assessing its impact on patient management and outcomes. Investigating the potential of the LT-VLA to predict other patient-centered outcomes, such as hospitalization rates and healthcare utilization, would provide a more comprehensive understanding of its utility.

## Conclusion

Our study demonstrates that improvements in the LT-VLA scale up to three years after lung transplant are associated with better HRQL, reduced risk of CLAD, and decreased mortality. The LT-VLA has the potential to be a prognostic tool in this unique patient population and offers a patient-centered approach to evaluating physical functioning and disability, complementing traditional assessment tools. Its integration into clinical practice and remote monitoring programs can enhance the management of lung transplant recipients, enabling timely interventions and potentially improving long-term outcomes. As healthcare continues to evolve towards more personalized and remote-based care, the LT-VLA scale may serve as one promising tool for advancing the evaluation and management of physical functioning in advanced lung disease and transplantation.

## Data Availability

All data produced in the present study are available upon reasonable request to the authors.

## Acknowledgments

The authors are deeply appreciative of the time our patient participants provided to support this research and members of the UCSF clinical Advanced Lung Disease and Transplant Program.

